# Pre-existing anxiety, depression, and neurological disability is associated with long COVID: A prospective and longitudinal cohort of the United Kingdom Multiple Sclerosis Register

**DOI:** 10.1101/2021.06.25.21259256

**Authors:** Afagh Garjani, Rodden M Middleton, Richard Nicholas, Nikos Evangelou

**Author notes:** Correspondence to Nikos Evangelou, Address: Department of Academic Clinical Neurology, C Floor, South Block, Queen’s Medical Centre, Nottingham NG7 2UH, United Kingdom.

## Abstract

**Objectives:** To assess the prevalence of long COVID among people with multiple sclerosis (MS) and its predictors, including their pre-COVID-19 functional status.

**Design:** Community-based prospective and longitudinal cohort study

**Setting:** The United Kingdom (UK) MS Register (UKMSR) COVID-19 study

**Participants:** A national cohort of people with MS and COVID-19

**Main outcome measures:** Participants used the online questionnaire-based platform of the UKMSR to update their COVID-19 symptoms, recovery status, and duration of symptoms for those who had fully recovered. Questionnaires were date-stamped for estimation of COVID-19 symptom duration for those who had not recovered at their last follow-up. The UKMSR holds demographic and up-to-date clinical data on participants as well as their web-based Expanded Disability Status Scale (a measure of physical disability in MS) and Hospital Anxiety and Depression Scale scores. The association between these factors and recovery from COVID-19 was assessed using multivariable Cox regression analysis.

**Results:** Out of 7,977 people with MS who participated in the UKMSR COVID-19 study, 599 had COVID-19 and updated their recovery status prospectively. At least 181 participants (31.1%) had long-standing COVID-19 symptoms for ≥4 weeks and 76 (13.1 %) for ≥12 weeks. Participants with higher levels of pre-COVID-19 physical disability, participants with anxiety and/or depression prior to COVID-19 onset, and women were less likely to report recovery from COVID-19.

**Conclusions:** Long COVID appears to disproportionately affect people with pre-existing mental health problems or physical disabilities. As post-COVID-19 rehabilitation services are being developed, individualised pathways should be considered to accommodate the needs of these vulnerable populations.

**Trial Registration:** ClinicalTrials.gov: NCT04354519

## INTRODUCTION

Long-term sequelae of COVID-19, or long COVID, has become a focus of research to inform the transformation of healthcare services for providing post-COVID-19 rehabilitation. Identification of long COVID risk factors will ensure that care is delivered more efficiently amid the burden of the COVID-19 pandemic on healthcare systems.^1^

A study has reported that asthma patients are at increased risk of experiencing long COVID.^2^ This finding may not be unexpected as respiratory symptoms constitute one of the most common symptoms of long COVID.^2 3^ It could be anticipated that people with other chronic conditions and symptoms similar to long COVID may also be vulnerable to the long-term effects of infection.

Neurological symptoms are common in long COVID with fatigue being the single most prevalent symptom.^2 3^ Fatigue is a prominent feature of many neurological conditions including multiple sclerosis (MS) with an estimated prevalence of 65.6%.^4^ Cognitive deficits, which are commonly reported in post-acute COVID-19,^5^ are estimated to affect 43-70% of people with MS.^6^ Similarly, studies have suggested a bidirectional association between psychiatric disorders and COVID-19 in that they can be both a risk factor for and a sequela of COVID-19.^7^

This study aimed to assess the prevalence and predictors of long COVID among people with MS.

## METHODS

This community-based prospective and longitudinal cohort study was conducted as part of the United Kingdom (UK) MS Register (UKMSR) COVID-19 study (ClinicalTrials.gov, Identifier: NCT04354519). Ethical approval for UKMSR studies was obtained from Southwest-Central Bristol Research Ethics Committee (16/SW/0194) and all participant provided informed consent, online.

People with MS had been reporting whether they had symptoms suggestive of COVID-19 from 17 March 2020-the start of the outbreak in the UK, using the platform of the UKMSR.^8^ Data were collected by online questionnaires. People with MS and COVID-19 (participants) were regularly reminded by email to update their COVID-19 symptoms and recovery status until reporting full recovery. Participants who reported full recovery also provided the duration of their COVID-19 symptoms. The submitted questionnaires were date-stamped for estimation of COVID-19 symptom duration for participants who had not reported full recovery at their last follow-up.

The UKMSR holds demographic and up-to-date clinical data on registered people with MS, including comorbidities, MS type, date of MS diagnosis, disease-modifying therapies, web-based Expanded Disability Status Scale (web-EDSS) scores, and Hospital Anxiety and Depression Scale (HADS) scores. The most recent web-EDSS and HADS scores before COVID-19 onset were used.

Web-EDSS measures neurological impairment in MS and is an ordinal scale, scored from 0 to 10, with higher scores indicating more physical disability.^9 10^ Participants were grouped into those with a web-EDSS score of (1) 0-2.5 (ambulatory without assistance-no or minimal neurological impairment), (2) 3-3.5 (ambulatory without assistance-moderate neurological impairment), (3) 4-5.5 (ambulatory without assistance-severe neurological impairment), (4) 6-6.5 (ambulatory with assistance), and (5) ≥7 (restricted to wheelchair or bed).

HADS is scored from 0 to 21 for anxiety and depression separately. Participants with scores ≥11 were considered as having probable anxiety or depression.^11^ Participants with anxiety, depression, or both were considered as one group (‘with anxiety and/or depression’) because these conditions frequently co-exist in people with MS,^12^ and the number of study participants with anxiety or depression alone was small.

Data collected until 19 March 2021 are presented according to STROBE guidelines.^13^

### Statistical Analysis

Data were analysed using IBM SPSS Statistics for Windows, version 26 (IBM Corp., Armonk, N.Y., USA; 2019).

Continuous variables with normal distribution are presented as mean (standard deviation [SD]) and were compared using the independent samples t-test. Continuous variables without normal distribution and ordinal variables are presented as median (interquartile range [IQR]) and were compared using the Mann-Whitney U test. The association between categorical variables was assessed using the Chi-square test or the Fisher’s exact test. The number of valid values for variables with missing data has been stated.

Univariable and multivariable Cox regression analysis, with time (days) from reporting COVID-19 to full recovery (‘event’) as the dependent variable, were performed to assess the association between demographic and clinical variables and recovery from COVID-19. Participants with persistent symptoms at their last follow-up were ‘censored’. A directed acyclic graph was produced (Supplemental Material) to identify potential confounding factors, which were subsequently accounted for in the multivariable Cox regression analysis. This method avoids the introduction of bias in the analysis by the erroneous inclusion of colliders and mediators as confounding factors.^14^ Listwise deletion was implemented for missing data. Results are presented as adjusted hazard ratios with 95% confidence intervals.

## RESULTS

Out of 7,977 people with MS who participated in the UKMSR COVID-19 study, 1,096 reported COVID-19. A total of 599 people with MS and COVID-19 updated their recovery status (participants) and 497 did not (non-participants). Participants did not differ in their baseline characteristics from non-participants (Table 1). Twenty-four participants had been hospitalised during their acute infection, 12 because of COVID-19 and 12 for non-COVID-19 reasons.

**Table 1.** Characteristics of people with MS and COVID-19 who updated (participants) or did not update (non-participants) their recovery status.

|  | Participants<br>n=599 | Participants with<br>confirmed COVID-19<sup>a</sup><br>n=208 | Non-participants<br>n=497 | p value<sup>b</sup> |
| --- | --- | --- | --- | --- |
| Age, mean (SD), years | 49 (11) | 49 (11) | 48 (11) | 0.212 |
| Women, n (%) | 462 (77.1) | 160 (76.9) | 395 (79.5) | 0.286 |
| White ethnicity, n (%) | 568 (94.8) | 199 (95.7) | 461 (92.8) | 0.155 |
| Comorbidities<sup>c</sup>, n (%) |  |  |  |  |
| Diabetes | 19 (4.1)<br>n=463 | 6 (3.9)<br>n=152 | 9 (2.7)<br>n=337 | 0.276 |
| Heart disease | 9 (1.9)<br>n=463 | 3 (2)<br>n=152 | 5 (1.5)<br>n=337 | 0.624 |
| Hyperlipidemia | 31 (6.7)<br>n=463 | 9 (5.9)<br>n=152 | 24 (7.1)<br>n=337 | 0.814 |
| Hypertension | 51 (11)<br>n=463 | 13 (8.6)<br>n=152 | 56 (16.6)<br>n=337 | 0.022 |
| Peripheral vascular disease | 1 (0.2)<br>n=463 | 0 (0)<br>n=152 | 0 (0)<br>n=337 | Not<br>applicable |
| Kidney disease | 7 (1.5)<br>n=463 | 2 (1.3)<br>n=152 | 5 (1.5)<br>n=337 | 0.974 |
| Liver disease | 2 (0.4)<br>n=463 | 1 (0.4)<br>n=152 | 2 (0.6)<br>n=337 | Not applicable |
| Lung disease | 55 (11.9)<br>n=463 | 13 (8.6)<br>n=152 | 54 (16)<br>n=337 | 0.092 |
| Anxiety and/or Depression<sup>d</sup> | 156 (38.5)<br>n=405 | 47 (33.8)<br>n=139 | 121 (37.7)<br>n=321 | 0.820 |
| Web-EDSS score<sup>c</sup>, median (IQR) | 4.5 (3-6.5)<br>n=421 | 4 (3-6.5)<br>n=147 | 4 (3-6.5)<br>n=293 | 0.886 |
| Web-EDSS score of 0 to 2.5, n (%) | 96 (22.8) | 30 (20.4) | 71 (24.2) | 0.905 |
| Web-EDSS score of 3 or 3.5, n (%) | 63 (15) | 25 (17) | 40 (13.7) |  |
| Web-EDSS score of 4 to 5.5, n (%) | 112 (16.6) | 42 (28.6) | 73 (24.9) |  |
| Web-EDSS score of 6 or 6.5, n (%) | 95 (22.6) | 29 (19.7) | 73 (24.9) |  |
| Web-EDSS score of $\geq 7$, n (%) | 55 (13.1) | 21 (14.3) | 36 (12.3) | |
| MS disease duration, median (IQR), years | 10 (5-17)<br>n=581 | 9 (4-15)<br>n=199 | 9 (5-16)<br>n=473 | 0.157 |
| Type of MS, n (%) |  |  |  |  |
| RRMS | 424 (70.8) | 152 (73.1) | 356 (71.6) | 0.403 |
| SPMS | 109 (18.2) | 33 (15.9) | 78 (15.7) |  |
| PPMS | 40 (6.7) | 15 (7.2) | 32 (6.4) |  |
| Unknown | 26 (4.3) | 8 (3.8) | 31 (6.2) |  |
| Taking a DMT, n (%) | 303 (50.6) | 107 (51.4) | 264 (53.1) | 0.403 |
| DMT = disease-modifying therapy; IQR = interquartile range; MS = Multiple sclerosis; PPMS = primary progressive MS; RRMS = Relapsing-remitting MS; SPMS = secondary progressive MS; Web-EDSS = web-based Expanded Disability Status Scale<br><sup>a</sup> A diagnosis of COVID-19 confirmed by a healthcare provider or testing.<br><sup>b</sup> Comparisons were made between all participants and non-participants.<br><sup>c</sup> Prior to COVID-19 onset.<br><sup>d</sup> Patients with Hospital Anxiety and Depression Scale scores $\geq 11$ for anxiety or depression were considered as having probable anxiety or depression, respectively. | | | | |

Four hundred and fifty-eight participants (76.5%) reported full recovery from COVID-19 at their last follow-up. Their median (IQR) symptom duration was 10 (6-21) days (n=455); 75 recovered in ≥4 weeks and 9 in ≥12 weeks. However, 141 participants (23.5%) had persistent symptoms at their last follow-up. They had been followed up for a median (IQR) of 91 (42-196) days (n=127) with 106 having symptoms for ≥4 weeks and 67 for ≥12 weeks from reporting COVID-19. Therefore, at least 181 participants (31.1%) had lasting COVID-19 symptoms for ≥4 weeks and 76 (13.1 %) for ≥12 weeks. The characteristics of participants by their symptom duration are compared in Table 2.

**Table 2.**
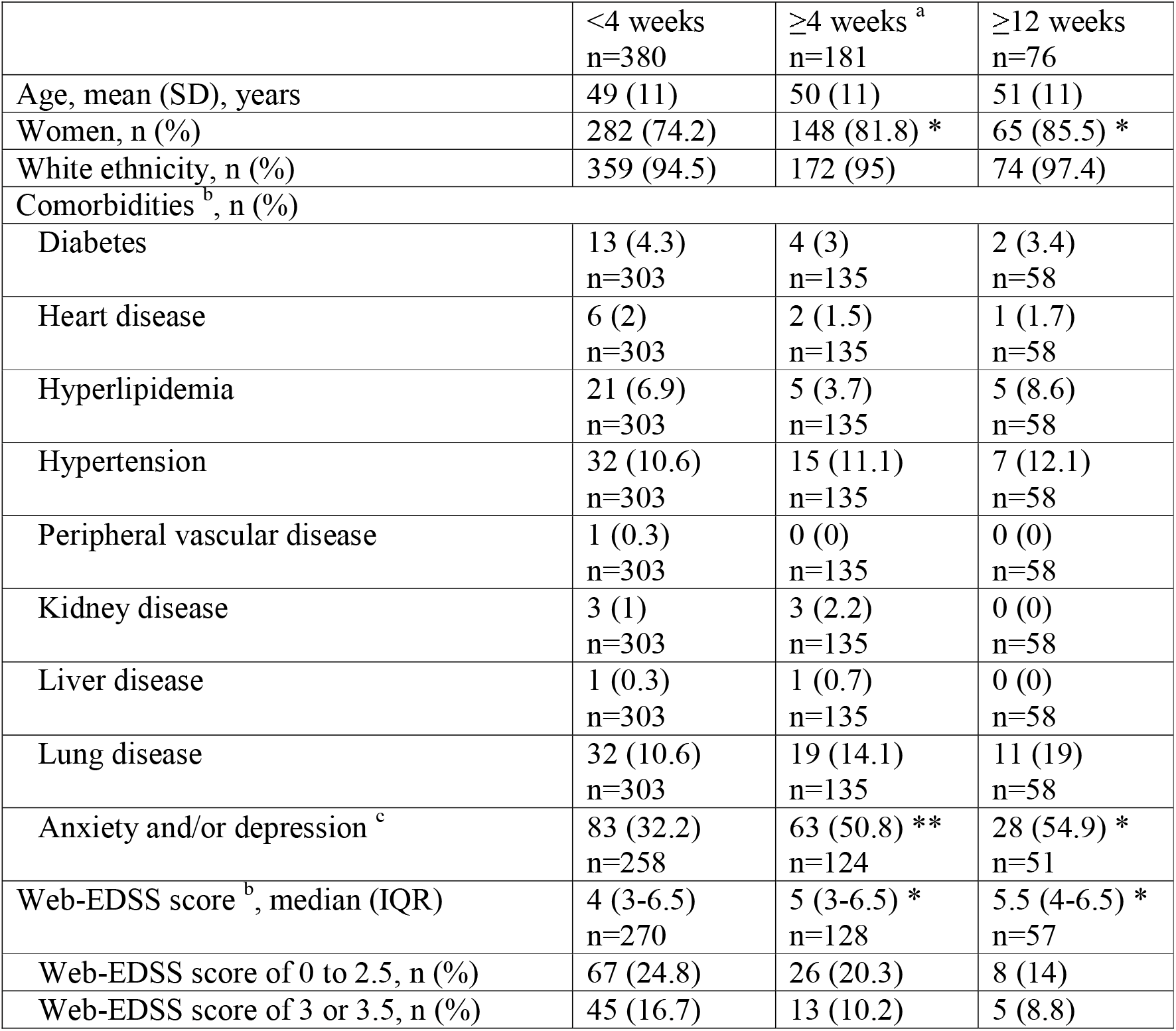
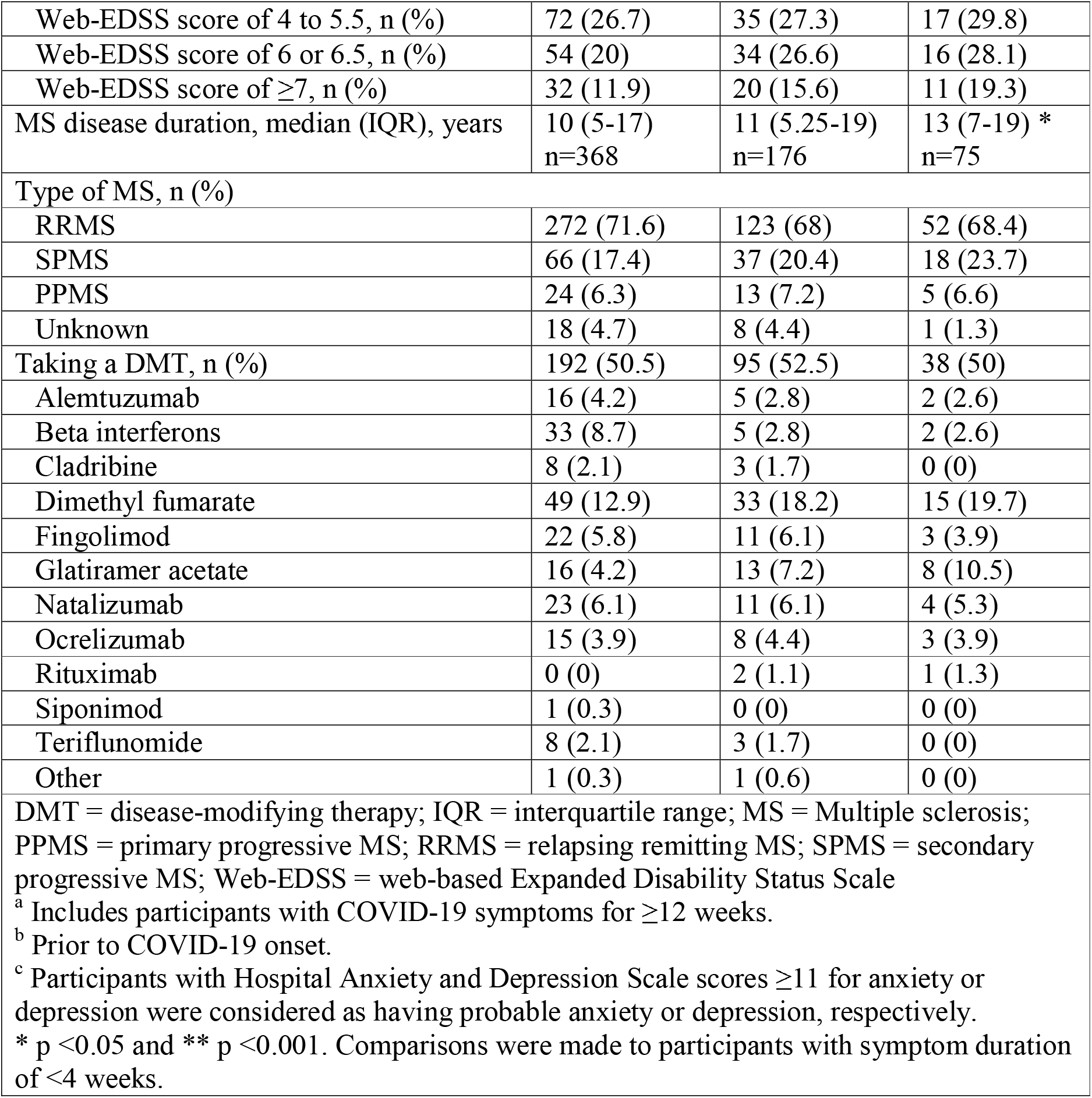
Characteristics of people with MS and COVID-19 in relation to the duration of their COVID-19 symptoms.

A post hoc analysis among participants with a COVID-19 diagnosis confirmed by a healthcare provider or testing showed similar findings (Supplemental Material). Participants with self-reported COVID-19 did not differ in their baseline characteristics from participants with confirmed COVID-19 (Table 1).

Participants with higher web-EDSS scores or anxiety and/or depression before COVID-19 onset and women were less likely to recover (Table 3).

**Table 3.**
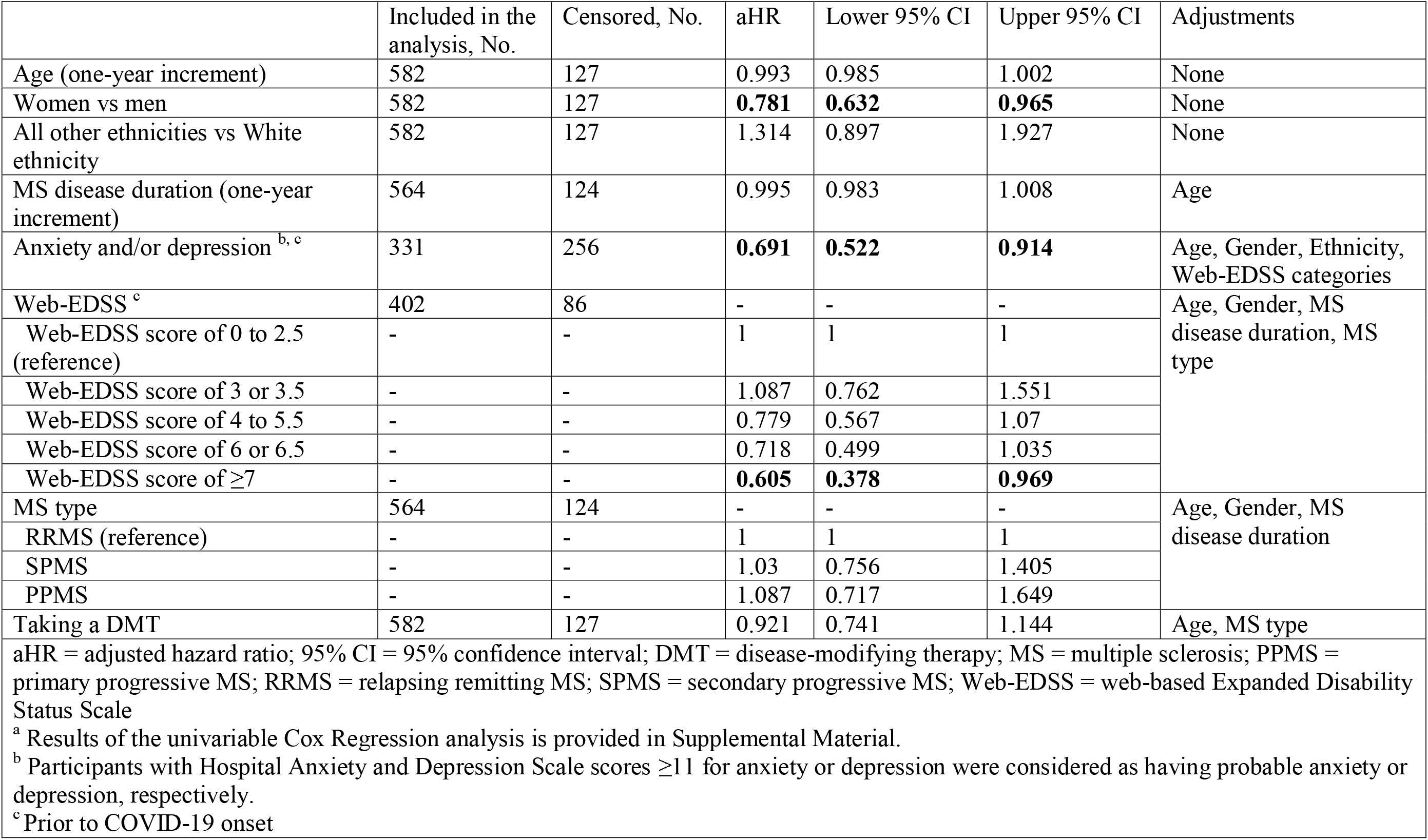
Results of the multivariable Cox regression analysis ^a^ of pre-COVID-19 factors associated with recovery from COVID-19.

New or worsening fatigue was the most common long COVID symptom followed by lower respiratory tract symptoms (Figure 1).

**Figure 1.**
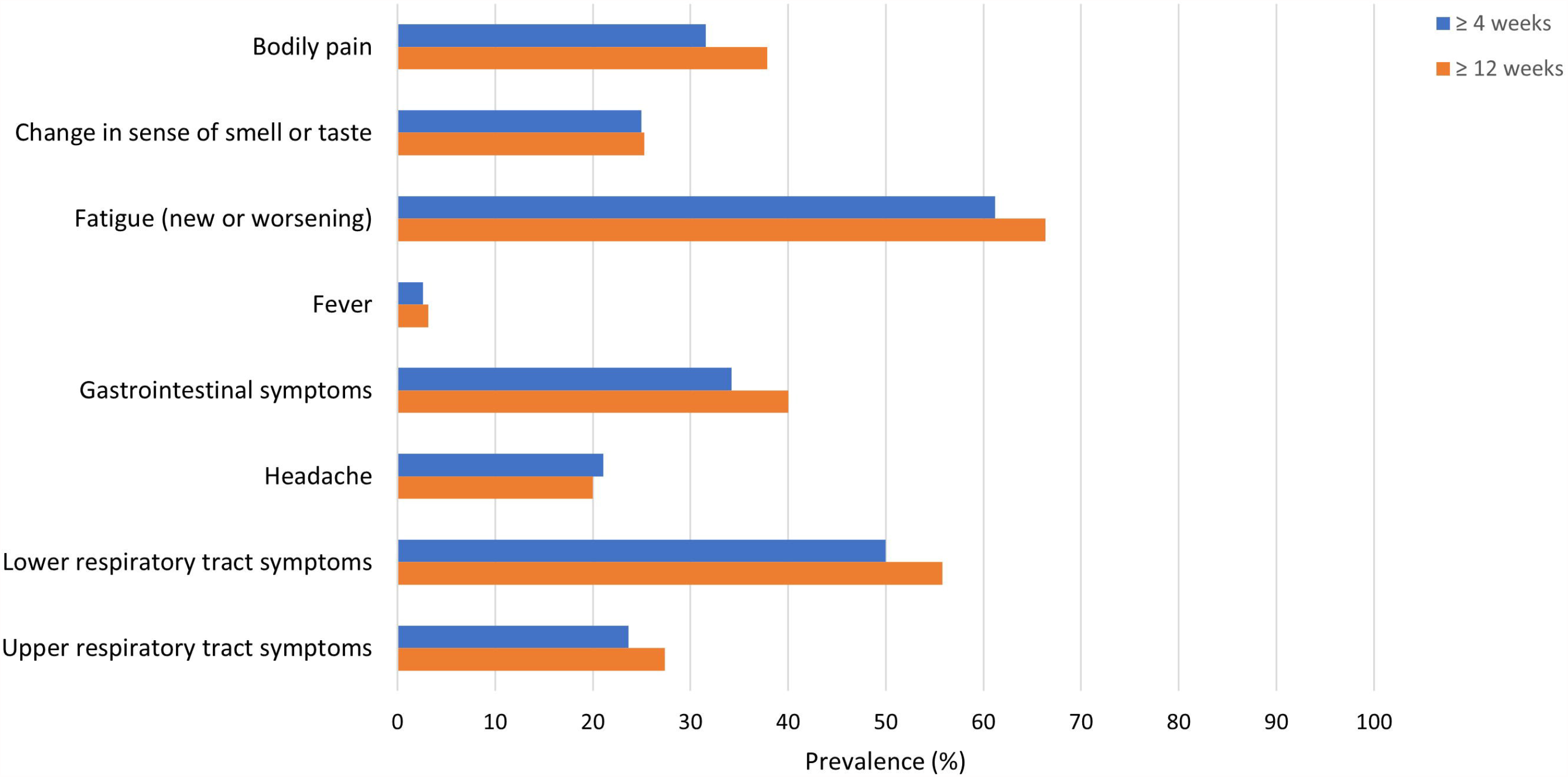
Frequency of COVID-19 symptoms among people with MS and COVID-19 who have had persistent symptoms at their last follow-up in ≥4 and ≥12 weeks from reporting COVID-19. Gastrointestinal symptoms included diarrhoea, nausea or vomiting, or stomach pain. Lower respiratory tract symptoms included coughs, shortness of breath, or heaviness in the chest. Upper respiratory tract symptoms included sore throat, nasal congestion, or sneezing.

## DISCUSSION

Studies have suggested that pre-existing neurological and psychiatric disorders are predisposing factors for COVID-19.^7^ In this prospective and longitudinal study, we report an association between pre-existing anxiety and depression and long COVID. Although the results of this study are from a population with higher prevalence of anxiety and depression than the general population,^15^ they highlight that patients with pre-COVID-19 mental health problems could be disproportionately affected by long COVID. The links between psychological stress and the function of the immune system could be one explanation for this observation.^16^

We also report a higher rate of long COVID in people with MS compared to a study of the general population with a similar methodology, using prospective data collection (2.3% for ≥12 weeks).^2^ Other studies are emerging that report much higher rates of long COVID (38% for ≥12 weeks) in the general population, but they have used retrospective data collection that can be affected by recall bias.^17^ Although the lack of a direct control group, for example recording any new symptoms in people with MS without COVID-19, is a limitation of this study, we feel that it is unlikely to affect these results. These findings in a study population comprised of mostly non-hospitalised people with MS and COVID-19 highlight previous observations that people with COVID-19 treated in the community can also experience prolonged symptoms of COVID-19.^2 18^

An association between pre-COVID-19 physical disability in people with MS and adverse acute COVID-19 outcomes has been previously reported.^19^ Our study shows that higher levels of physical disability predispose them to long COVID as well.

It is possible that long-lasting symptoms of COVID-19 in populations with asthma or MS be a deterioration of their pre-existing condition triggered by the infection rather than persistent new symptoms of COVID-19? It is known that respiratory infections can trigger asthma exacerbations,^20^ and we have previously shown that COVID-19 can lead to MS exacerbations.^21^ New or worsening fatigue was the most common persisting symptom in our population followed by lower respiratory tract symptoms, a pattern similar to the general population.^2 3^ Fatigue, however, is also prevalent in MS.^4^ Further research is needed to understand what is long COVID and whether it presents with different symptom patterns in populations with other pre-existing disorders.

We think these observations are important as healthcare services are being reformed to address the consequences of the pandemic. This study adds to studies of long COVID in the general population,^2 22^ highlighting that, in long-term, COVID-19 could affect people with pre-existing conditions differently. These findings have implications for long COVID research-to be inclusive of at-higher risk populations, and for provision of post-COVID-19 rehabilitation-as individualized pathways to accommodate the needs of these potentially more vulnerable populations are needed.

## Data Availability

Data are stored on the UKMSR Secure e-Research Platform at Swansea University Medical School. Line level data cannot be released, but qualified researchers, subject to governance, can request access to data.

## Acknowledgments

None.

## REFERENCES

1. Maxwell E. Unpacking post-covid symptoms: British Medical Journal Publishing Group, 2021.

2. Sudre CH, Murray B, Varsavsky T, et al. Attributes and predictors of long COVID. Nature Medicine 2021;27(4):626–31.

3. Logue JK, Franko NM, McCulloch DJ, et al. Sequelae in adults at 6 months after COVID-19 infection. JAMA network open 2021;4(2):e210830–e30.

4. Weiland TJ, Jelinek GA, Marck CH, et al. Clinically significant fatigue: prevalence and associated factors in an international sample of adults with multiple sclerosis recruited via the internet. PLoS One 2015;10(2):e0115541.

5. Hampshire A, Trender W, Chamberlain S, et al. Cognitive deficits in people who have recovered from COVID-19 relative to controls: An N= 84,285 online study. MedRxiv 2020

6. Chiaravalloti ND, DeLuca J. Cognitive impairment in multiple sclerosis. The Lancet Neurology 2008;7(12):1139–51.

7. Taquet M, Geddes JR, Husain M, et al. 6-month neurological and psychiatric outcomes in 236 379 survivors of COVID-19: a retrospective cohort study using electronic health records. The Lancet Psychiatry 2021

8. Evangelou N, Garjani A, Hunter R, et al. Self-diagnosed COVID-19 in people with multiple sclerosis: a community-based cohort of the UK MS Register. Journal of Neurology, Neurosurgery & Psychiatry 2021;92(1):107–09.

9. Kurtzke JF. Rating neurologic impairment in multiple sclerosis: an expanded disability status scale (EDSS). Neurology 1983;33(11):1444–44.

10. Leddy S, Hadavi S, McCarren A, et al. Validating a novel web-based method to capture disease progression outcomes in multiple sclerosis. Journal of neurology 2013;260(10):2505–10.

11. Marrie RA, Zhang L, Lix LM, et al. The validity and reliability of screening measures for depression and anxiety disorders in multiple sclerosis. Multiple sclerosis and related disorders 2018;20:9–15.

12. Wood B, Van Der Mei I, Ponsonby A-L, et al. Prevalence and concurrence of anxiety, depression and fatigue over time in multiple sclerosis. Multiple Sclerosis Journal 2013;19(2):217–24.

13. Von Elm E, Altman DG, Egger M, et al. The Strengthening the Reporting of Observational Studies in Epidemiology (STROBE) statement: guidelines for reporting observational studies. Annals of internal medicine 2007;147(8):573–77.

14. Williams TC, Bach CC, Matthiesen NB, et al. Directed acyclic graphs: a tool for causal studies in paediatrics. Pediatric research 2018;84(4):487–93.

15. Marrie RA, Walld R, Bolton JM, et al. Estimating annual prevalence of depression and anxiety disorder in multiple sclerosis using administrative data. BMC research notes 2017;10(1):619.

16. Segerstrom SC, Miller GE. Psychological stress and the human immune system: a meta-analytic study of 30 years of inquiry. Psychological bulletin 2004;130(4):601.

17. Matthew W, Joshua E, Marc C-H, et al. Persistent symptoms following SARS-CoV-2 infection in a random community sample of 508,707 people 2021 [Available from: https://spiral.imperial.ac.uk/handle/10044/1/89844 accessed 24 June 2021.

18. Tenforde MW, Kim SS, Lindsell CJ, et al. Symptom duration and risk factors for delayed return to usual health among outpatients with COVID-19 in a multistate health care systems network—United States, March–June 2020. Morbidity and Mortality Weekly Report 2020;69(30):993.

19. Salter A, Fox RJ, Newsome SD, et al. Outcomes and Risk Factors Associated With SARS-CoV-2 Infection in a North American Registry of Patients With Multiple Sclerosis. JAMA neurology 2021

20. Singh A, Busse W. Asthma exacerbations· 2: Aetiology. Thorax 2006;61(9):809–16.

21. Garjani A, Middleton RM, Hunter R, et al. COVID-19 is associated with new symptoms of multiple sclerosis that are prevented by disease modifying therapies. Multiple Sclerosis and Related Disorders 2021.

22. Daugherty SE, Guo Y, Heath K, et al. Risk of clinical sequelae after the acute phase of SARS-CoV-2 infection: retrospective cohort study. bmj 2021;373

