## Supplemental Material for "Pre-existing anxiety, depression, and neurological disability is associated with long COVID: A prospective and longitudinal cohort of the United Kingdom Multiple Sclerosis Register"

**Methods**

A directed acyclic graph (DAG; Supplemental Material-Figure 1) was used to identify potential confounding factors for inclusion in the multivariable Cox regression analysis.

**Supplemental Material-Figure 1.** The directed acyclic graph used for identifying confounding factors.

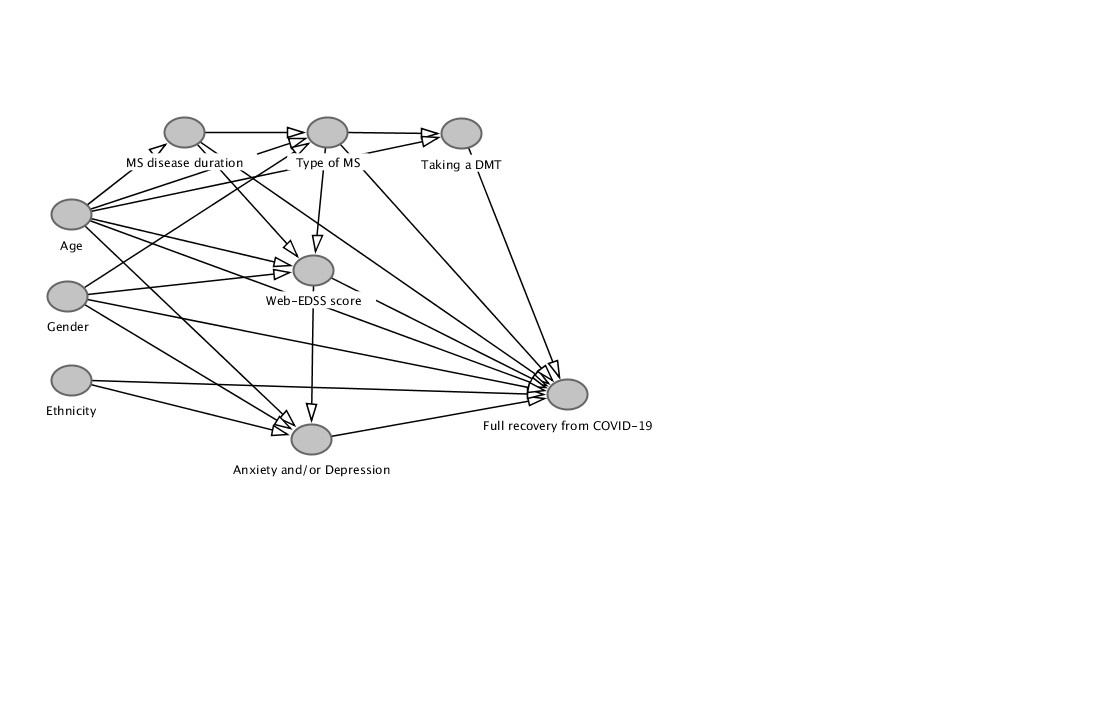

The following code can be used at “http://www.dagitty.net/dags.html” to replicate the DAG:

dag {

bb="0,0,1,1"

"Anxiety and/or Depression" [pos="0.284,0.611"]

"Full recovery from COVID-19" [pos="0.518,0.549"]

"MS disease duration" [pos="0.168,0.184"]

"Taking a DMT" [pos="0.421,0.186"]

"Type of MS" [pos="0.299,0.184"]

"Web-EDSS score" [pos="0.286,0.376"]

Age [pos="0.065,0.299"]

Ethnicity [pos="0.065,0.529"]

Gender [pos="0.062,0.412"]

"Anxiety and/or Depression" -> "Full recovery from COVID-19"

"MS disease duration" -> "Full recovery from COVID-19"

"MS disease duration" -> "Type of MS"

"MS disease duration" -> "Web-EDSS score"

"Taking a DMT" -> "Full recovery from COVID-19"

"Type of MS" -> "Full recovery from COVID-19"

"Type of MS" -> "Taking a DMT"

"Type of MS" -> "Web-EDSS score"

"Web-EDSS score" -> "Anxiety and/or Depression"

"Web-EDSS score" -> "Full recovery from COVID-19"

Age -> "Anxiety and/or Depression"

Age -> "Full recovery from COVID-19"

Age -> "MS disease duration"

Age -> "Taking a DMT"

Age -> "Type of MS"

Age -> "Web-EDSS score"

Ethnicity -> "Anxiety and/or Depression"

Ethnicity -> "Full recovery from COVID-19"

Gender -> "Anxiety and/or Depression"

Gender -> "Full recovery from COVID-19"

Gender -> "Type of MS"

Gender -> "Web-EDSS score"

}

**Results**

*COVID-19 symptom duration among participants with confirmed COVID-19*

In the UK, mass testing for COVID-19 was implemented on 28 May 2020. Three hundred and fifteen participants (52.6%) had had COVID-19 before this date, when they could not have been tested outside of hospitals. A total of 208 participants (34.7%) had their diagnosis confirmed by a healthcare provider or testing. A hundred and thirty-eight participants with confirmed COVID-19 (66.3%) reported full recovery. Their median (IQR) symptom duration was 12 (7–21) days (n=137) with 23 experiencing symptoms for ≥4 weeks and 2 for ≥12 weeks. Participants with confirmed COVID-19 and persistent symptoms at their last follow-up had been followed up for a median (IQR) of 52 (37-185) days (n=63) with 51 having lasting symptoms for ≥4 weeks and 25 for ≥12 weeks. As a result, at least 37% of participants with confirmed COVID-19 (n=74) had lasting COVID-19 symptoms for ≥4 weeks and 13.5% (n=27) for ≥12 weeks.

**Supplemental Material Table 1.** Results of the univariable Cox regression analysis of pre-COVID-19 factors associated with recovery from COVID-19.

|  | Included in the analysis, No. | Censored, No. | HR | Lower 95% CI | Upper 95% CI |
| --- | --- | --- | --- | --- | --- |
| Age (one-year increment) | 582 | 127 | 0.993 | 0.985 | 1.002 |
| Women vs men | 582 | 127 | **0.781** | **0.632** | **0.965** |
| All other ethnicities vs White ethnicity | 582 | 127 | 1.314 | 0.897 | 1.927 |
| MS disease duration (one-year increment) | 564 | 124 | 0.992 | 0.982 | 1.002 |
| Anxiety and/or depression ^a, b^ | 395 | 89 | **0.680** | **0.536** | **0.862** |
| Web-EDSS ^b^ | 412 | 88 | - | - | - |
| Web-EDSS score of 0 to 2.5 (reference) | - | - | 1 | 1 | 1 |
| Web-EDSS score of 3 or 3.5 | - | - | 1.037 | 0.731 | 1.473 |
| Web-EDSS score of 4 to 5.5 | - | - | 0.829 | 0.612 | 1.123 |
| Web-EDSS score of 6 or 6.5 | - | - | 0.748 | 0.542 | 1.032 |
| Web-EDSS score of ≥7 | - | - | 0.722 | 0.489 | 1.066 |
| MS type | 582 | 127 | - | - | - |
| RRMS (reference) | - | - | 1 | 1 | 1 |
| SPMS | - | - | 0.920 | 0.718 | 1.178 |
| PPMS | - | - | 1.054 | 0.716 | 1.550 |
| Taking a DMT | 582 | 127 | 0.964 | 0.802 | 1.158 |
| HR = unadjusted hazard ratio; 95% CI = 95% confidence interval; DMT = disease-modifying therapy; MS = multiple sclerosis; PPMS = primary progressive MS; RRMS = relapsing remitting MS; SPMS = secondary progressive MS; Web-EDSS = web-based Expanded Disability Status Scale  ^a^ Participants with Hospital Anxiety and Depression Scale scores ≥11 for anxiety or depression were considered as having probable anxiety or depression, respectively.  ^b^ Prior to COVID-19 onset | | | | | |
